# Reduced seroconversion in children compared to adults with mild COVID-19

**DOI:** 10.1101/2021.10.17.21265121

**Authors:** Zheng Quan Toh, Jeremy Anderson, Nadia Mazarakis, Melanie Neeland, Rachel A. Higgins, Karin Rautenbacher, Kate Dohle, Jill Nguyen, Isabella Overmars, Celeste Donato, Sohinee Sarkar, Vanessa Clifford, Andrew Daley, Suellen Nicholson, Francesca L. Mordant, Kanta Subbarao, David P. Burgner, Nigel Curtis, Julie E. Bines, Sarah McNab, Andrew C. Steer, Kim Mulholland, Shidan Tosif, Nigel W. Crawford, Daniel G. Pellicci, Lien Anh Ha Do, Paul V Licciardi

## Abstract

**Importance:** The immune response in children with SARS-CoV-2 infection is not well understood.

**Objective:** To compare seroconversion in children and adults with non-hospitalized (mild) SARS-CoV-2 infection and to understand the factors that influence this.

**Design:** Participants were part of a household cohort study of SARS-CoV-2 infection. Weekly nasopharyngeal/throat swabs and blood samples were collected during the acute and convalescent period following PCR diagnosis for analysis.

**Setting:** Participants were recruited at the Royal Children’s Hospital, Melbourne, Australia between May and October 2020.

**Participants:** Those who had a SARS-CoV-2 PCR-positive nasal/throat swab.

**Main outcomes and measures:** SARS-CoV-2 antibody and cellular responses in children and adults. Seroconversion was defined by seropositivity in all three serological assays.

**Results:** Among 108 SARS-CoV-2 PCR-positive participants, 57 were children (median age: 4, IQR 2-10) and 51 were adults (median age: 37, IQR 34-45). Using three established serological assays, a lower proportion of children seroconverted compared with adults [20/54 (37.0%) vs 32/42 (76.2%); (p<0.001)]. This was not related to viral load, which was similar in children and adults [mean Ct 28.58 (SD: 6.83) vs 24.14 (SD: 8.47)]. Age and sex also did not influence seroconversion or the magnitude of antibody response within children or adults. Notably, in adults (but not children) symptomatic adults had three-fold higher antibody levels than asymptomatic adults (median 227.5 IU/mL, IQR 133.7-521.6 vs median 75.3 IU/mL, IQR 36.9-113.6). Evidence of cellular immunity was observed in adults who seroconverted but not in children who seroconverted.

**Conclusion and Relevance:** In this non-hospitalized cohort with mild COVID-19, children were less likely to seroconvert than adults despite similar viral loads. This has implications for future protection following COVID-19 infection in children and for interpretation of serosurveys that involve children. Further research to understand why children are less likely to seroconvert and develop symptoms following SARS-CoV-2 infection, and comparison with vaccine responses may be of clinical and scientific importance.

**Key points:** *Question:* What proportion of children with non-hospitalized (mild) SARS-CoV-2 infection seroconvert compared to adults?

*Findings:* In this cohort study conducted in 2020, we found the proportion of children who seroconverted to SARS-CoV-2 was half that in adults despite similar viral load.

*Meaning:* Serology is a less reliable marker of prior SARS-CoV-2 infection in children. SARS-CoV-2-infected children who do not seroconvert may be susceptible to reinfection. Our findings support strategies to protect children against COVID-19 including vaccination.

## Introduction

Since the start of the COVID-19 pandemic, most children with COVID-19 have been either asymptomatic or present with mild illness, and very few require hospitalization^1-3^. However, COVID-19 cases in children are increasing in 2021 due to the emergent of SARS-CoV-2 variants, particularly the Delta variant, posing important questions regarding the immune responses in children^4,5^. While the severity of COVID-19 generally correlates with the magnitude of host immune responses against SARS-CoV-2^6,7^, children and adolescents with mild or asymptomatic SARS-CoV-2 infection can also mount robust and durable antibody responses^8^.

Immunity to SARS-CoV-2 induced through natural infection is likely to be mediated by a combination of humoral and cellular immunity^9-11^. Some studies comparing children and adults have revealed distinct immune profiles^12-15^, which have been associated with less severe outcomes in children compared with adults.

The correlates of protection against SARS-CoV-2 have not been identified, although neutralizing antibodies are increasingly recognized as the primary mediator of protection^16-18^. The majority of adults (>90%) infected with SARS-CoV-2 mount an antibody response^19,20^, which can persist for at least 12 months^21^. Seropositive recovered adults are estimated to have up to 89% protection from reinfection against the same strain^22,23^. In contrast, the proportion of children infected with SARS-CoV-2 who seroconvert is unknown, particularly among those with asymptomatic or mild infection.

Characterization of the immune response following natural infection is important to better understand factors that may be related to future protection. In this study, we compared seroconversion and cellular immunity in children and adults following infection with the ancestral (Wuhan) strain of SARS-CoV-2 and investigated the factors associated with this response in a household cohort study in Melbourne, Australia.

## Methods

### Study cohort

Participants were recruited as part of a household cohort study at The Royal Children’s Hospital, Melbourne, Australia between May and October 2020. Infection with SARS-CoV-2 was confirmed by PCR of nasopharyngeal/oropharyngeal (NP) swabs. Participants were non-hospitalized patients who were asymptomatic or had mild symptoms of COVID-19 (i.e. coryza, headaches, nausea, fever, cough, sore throat, malaise and/or muscle aches). Baseline swab and convalescent blood samples (median 41 days, interquartile range, IQR: 31-49) were collected from all individuals. A subset of individuals had two to four additional weekly NP swabs and a baseline blood sample collected (median 7-12 days, IQR: 4-13 following the baseline swab), as well as a later blood sample collected at median 94 days (IQR: 91-100). Written informed consent and assent were obtained from adults/parents and children, respectively. The study was done with the approval of the Human Research Ethics Committee (HREC) at the Royal Children’s Hospital, Melbourne: HREC/63666/RCHM-2019.

### SARS-CoV-2 PCR diagnosis

Combined oropharyngeal and nasopharyngeal (or deep nasal) swabs were collected using dry FLOQSwabs® (Copan, Brescia, Italy). Briefly, FLOQSwabs were eluted in phosphate buffered saline (PBS) and the eluent was used for nucleic acid extraction using the Roche MagNA Pure 96 extraction system (Roche, Basel, Switzerland), according to the manufacturer’s instructions. The majority of SARS-CoV-2 samples were initially tested using the LightMix® Modular SARS and Wuhan CoV E-gene kit (targeting the E-gene; sensitivity 96.5%, specificity of 98.5% ^24^; TIB Molbiol, Berlin, Germany) using 10 μL nucleic acid extract, according to the manufacturer’s instructions. RT-qPCR was performed on the LightCycler 480 II Real-Time PCR System (Roche). Ct values at diagnosis for SARS-CoV-2 positive patients are provided, when available.

### SARS-CoV-2 serology diagnosis

#### In-house ELISA method

We used a modified two-step ELISA based on the Mount Sinai Laboratory method previously described ^25,26^. Briefly, 96-well high-binding plates were coated with receptor binding domain (RBD) or S1 (Sino Biological, China) antigen diluted in PBS at 2 µg/mL. Serum samples were first screened with RBD antigen, and potential seropositive samples were then confirmed with S1 antigen. Goat anti-human IgG-(1:10,000) horseradish peroxidase (HRP) conjugated secondary antibody was used, and the plates were developed using 3.3’, 5.5’-tetramethylbenzidine substrate solution. Seropositive samples were titrated and calculated based on a World Health Organization (WHO) SARS-CoV-2 pooled serum standard (National Institute of Biological Standards and Controls, United Kingdom). Results were reported in International Units/mL. The cut-off for seropositivity was 8.36 IU/mL based on pre-pandemic samples, while seronegative samples were given half of the seropositive cut-off value.

#### Liaison SARS-CoV-2 S1/S2 IgG assay (Saluggia, Italy)

The quantitative commercial assay for the detection of IgG antibodies against S1/S2 antigens of SARS-CoV-2 was done according to the manufacturer’s instructions. Data was reported as Assay Units (AU)/mL; negative (<12.0 AU/mL), equivocal (12.0-15.0 AU/mL), or positive (>15.0 AU/mL).

#### Wantai SARS-CoV-2 antibody ELISA (Beijing, China)

This qualitative commercial assay detects total antibodies (including IgG and IgM) to SARS-CoV-2 RBD antigen. The assays were done according to the manufacturer’s instructions. Data was reported as a ratio of the absorbance over the kit control cut-off; seropositive is defined as ratio ≥ 1.0.

### Flow cytometry

For T- and B-cell populations (convalescent sample), whole blood was lysed with red blood cell lysis buffer (1:10 dilution) for 10 min at room temperature (RT). Whole blood was then diluted in PBS and centrifuged at 400 x g for 5 min. Cells were washed once more in PBS then resuspended in 50 µL blocking solution (1% FC-block and 5% normal rat serum in PBS) for 15 min on ice. Following blocking, cells were washed with 1 mL of FACS buffer (2% FBS in PBS) then stained with 50 µL of antibody cocktail 1 or 2 for 20 min on ice (Supplementary Table 1). After staining, cells were washed twice then resuspended in 100 µL of FACS buffer for acquisition using the Cytek Aurora. Compensation was performed at the time of acquisition using compensation beads (BD Bioscience, San Diego, CA, USA). Data were analysed by FlowJo (Tree Star). Supplementary Figures 1 and 2 depicts the manual gating strategy for B- and T cell panels.

For innate cell populations (baseline sample), 100 µL of whole blood was aliquoted for flow cytometry analysis. Whole blood was lysed with 1 mL of red cell lysis buffer for 10 min at room temperature. Cells were washed with 1mL PBS and centrifuged at 350 x g for 5 min. Following two more washes, cells were resuspended in PBS for viability staining using near infra-red viability dye according to manufacturer’s instructions. The viability dye reaction was stopped by the addition of FACS buffer (2% heat-inactivated FCS in 2 mM EDTA) and cells were centrifuged at 350 x g for 5 min. Cells were then resuspended in human FC-block for 5 min at room temperature. The whole blood innate cocktail (Supplementary Table 2) made up at 2X concentration were added 1:1 with the cells and incubated for 30 min on ice. Following staining, cells were washed with 2 mL FACS buffer and centrifuged at 350 x g for 5 min. Cells were then resuspended in 2% PFA for a 20 min fixation on ice, washed, and resuspended in 150µL FACS buffer for acquisition using the BD LSR X-20 Fortessa. Supplementary Figure 3 depicts the manual gating strategy for innate cell populations.

### Statistical analysis

The antibody levels and Ct values between children and adults, as well as within seropositive/seronegative children or adults were compared using Mann-Whitney U test. Fisher’s exact test was used to compare both the proportion who were seropositive and the proportion who were symptomatic in children and adults. For correlation analysis, antibody levels were log-transformed and analyzed using Pearson’s correlation analysis. All analyses were performed with GraphPad Prism 7.0. A p<0.05 was considered significant.

## Results

### Participants’ characteristics

Between May 10, 2020 and October 28, 2020, 134 children (less than 18 years of age) and 160 adults (19–73 years of age) from 95 families were recruited into the household cohort study. A total of 57/134 children (42.5%) and 51/160 adults (31.9%) were infected with SARS-CoV-2 (defined as having a positive PCR result for SARS-CoV-2 at any of the five timepoints) and were included in our analysis; 30/57 children and 19/51 adults had two to four additional weekly swabs collected. Four adults were PCR negative at baseline but returned a positive PCR result one week later. The median ages at enrolment for children was 4 years old (IQR: 2-10) and 37 years old for adults (IQR: 34-45). Among them, 22/57 (38.6%) children and 28/51 (54.9%) were female.

### Fewer children seroconvert compared to adults

Two commercial and one in-house assay were used to measure antibody responses in children and adults at acute (median day 7-12, IQR: 4-13) and convalescence timepoints (median day 41, IQR: 31-49). There was a significant increase in antibody levels between acute and convalescence for adults but not in children for all three assays (Fig. 1A). Results from all three assays were highly concordant, with 96/108 (88.9%) samples positive by all three assays (Fig. 1B, Supplementary Fig. 4), and 94-97% agreement between the assays (Supplementary Table 3). A subset of these samples was also tested by a SARS-CoV-2 microneutralization assay and the results positively correlated with results from all three assays (Supplementary Fig. 5). Interestingly, lower seropositivity was found in SARS-CoV-2-infected children (40.4-40.7%) at the convalescent timepoint compared to adults (61.4-73.7%, depending on the assay) (Fig. 1C). All seronegative children at the median convalescent timepoint (Day 41) did not become seropositive by the median Day 94 timepoint. (Fig. 1D).

**Fig. 1:**
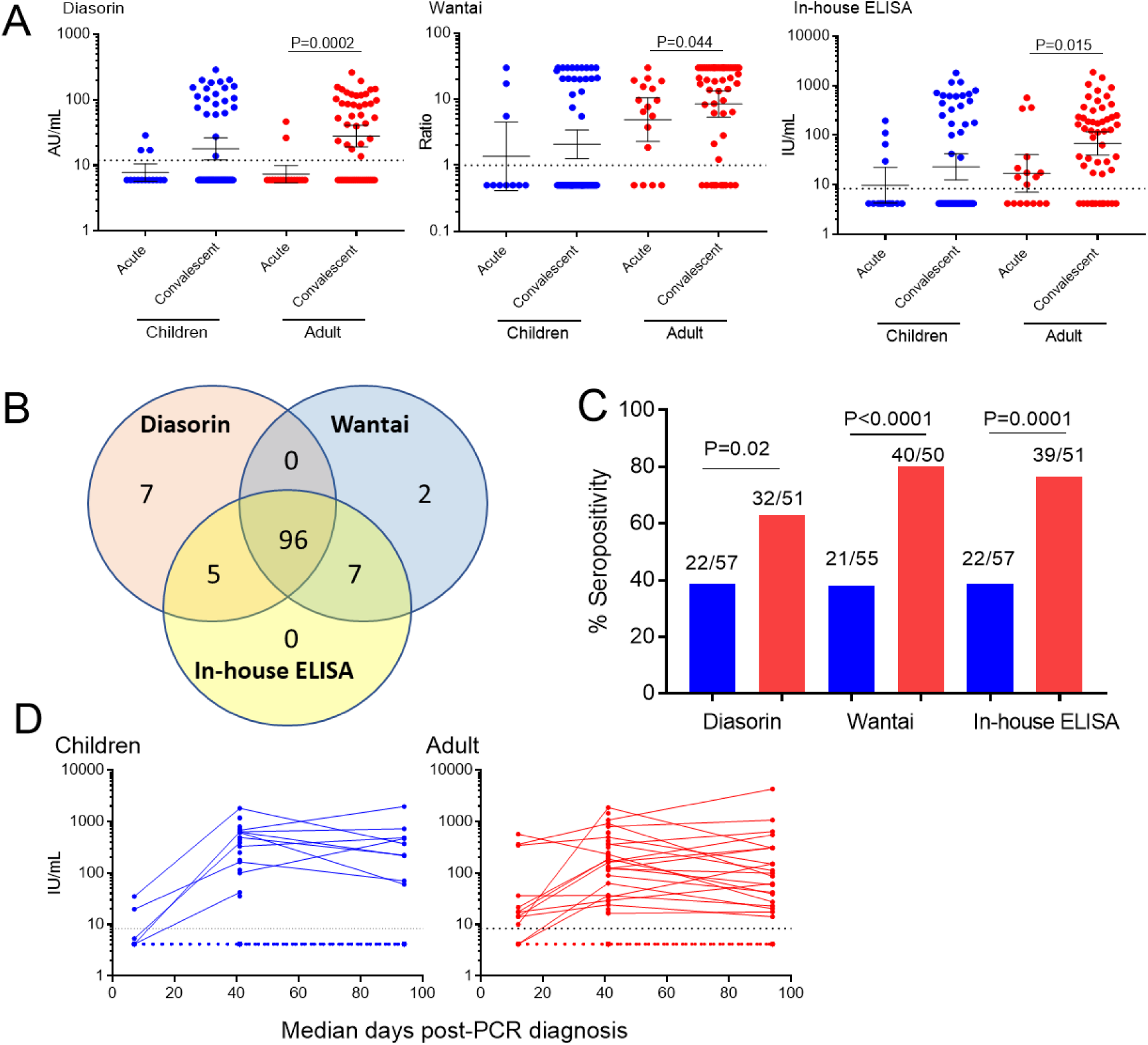
SARS-CoV-2 antibody response in children (blue) and adults (red) measured by three serological assays; Diasorin (S1/S2), Wantai (RBD) and in-house ELISA (RBD/S1). (A) Mean IgG SARS-CoV-2 antibody levels (±95%CI) at acute (median day 7-12, IQR: 4-13. Children, N=14; Adult, N=17) and convalescent (median day 41, IQR: 31-49. Children, N=57; Adult, N=51). (B) Venn diagram showing the concordance of seropositivity results between the three serological assays (N=105-108); two children and 1 adult sample were not tested by Wantai assay. (C) Seropositivity rate in children and adults at convalescent period (median day 41. Children, N=57; Adult, N=51). (D) SARS-CoV-2 IgG levels over time in children and adults using an in-house ELISA; number of samples per timepoint: Children, Day 7 (N=13), Day 41 (N=59), Day 94 (N=26); Adult, Day 12 (N=20), Day 41 (N=57), Day 94 (N=29).

### Factors that may influence the antibody response

To investigate the factors involved in seroconversion, we included only those participants who were seropositive or seronegative by all three serological assays (Fig. 2A); 9 samples from adults and 3 from children were excluded due to inconsistent serostatus or not tested on all assays due to sample availability. We found no difference in viral loads at baseline between children and adults [mean Ct 28.58 (SD: 6.83) vs mean Ct 24.14 (SD: 8.47)] (Fig. 2B). The time between PCR diagnosis to convalescent sampling was also similar (median days 41, IQR, 31-49 vs median days 41, IQR, 35-49) (Fig 2C).

**Fig. 2:**
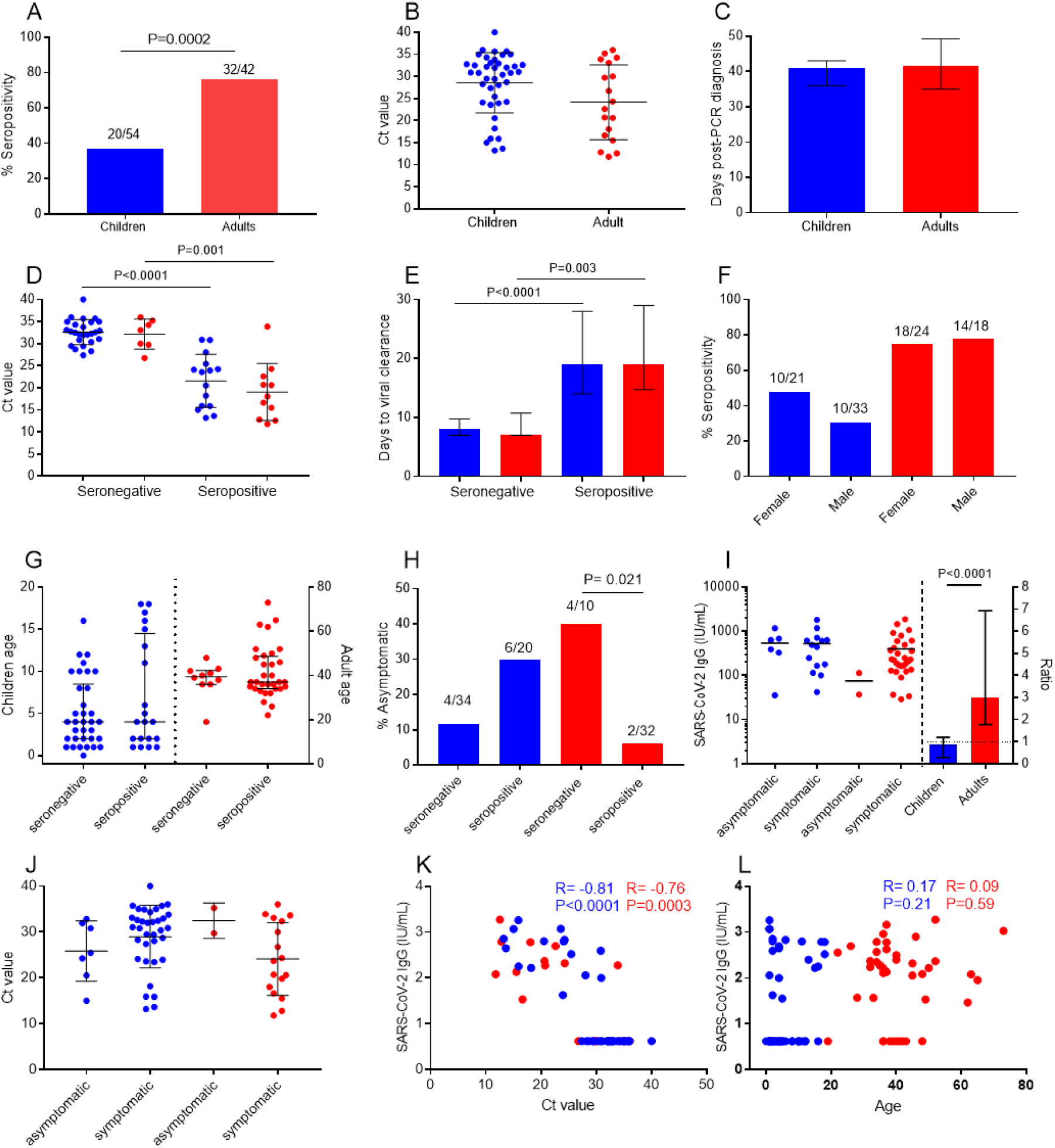
Factors associated with SARS-CoV-2 antibody responses based on in-house ELISA assay. (A) Seropositivity rate in children (blue) and adults (red) at convalescent period (median day 41. Children, N=54; Adult, N=42) who were seropositive and seronegative by all three serological assays. (B) Mean viral load (SD) between children (N=42) and adult (N=18) where data were available. (C) Median days (IQR) between positive PCR diagnosis and convalescent blood sampling between children (N=54) and adult (N=42). (D) Mean viral load (SD) between children (N=42) and adult (N=18) stratified by serostatus. (E) Duration of viral clearance (median days, IQR) stratified by serostatus (seronegative children, N=20, seropositive children, N=7; seronegative adult, N=4, seropositive adult, N=10). (F) Seropositivity rate stratified by sex. (G) Age of children and adults stratified by serostatus (Children, N=54; Adult, N=42) (Median, IQR). (H) Proportion of asymptomatic children and adults stratified by serostatus. (I) Median antibody levels (IQR) based on symptoms (left y-axis) and median fold-change in antibody levels between asymptomatic and symptomatic in children (N=6 vs N=14) and adults (N=2 vs N=30) (right y-axis). (J) Mean viral load (SD) stratified based on symptoms in children (asymptomatic, N=7 vs symptomatic, N=35) and adults (asymptomatic, N=2 vs symptomatic, N=17). (K) Correlation between antibody levels and viral load. (L) Correlation between antibody levels and age. Blue dots/bars represent children and red dots/bars represent adults. Seropositivity was defined as seropositive by all three assays. Pearson’s correlation analysis was used to examine association. Ct value: cycle threshold.

Individuals were more likely to be seropositive with higher viral loads and longer viral clearance time (based on those with multiple swabs collected), but there were no differences in these parameters between children and adults who were seronegative or seropositive (Fig 2D-E). Interestingly, a Ct value of less than 26 was associated with seroconversion in 80% (12/15) and 91% (10/11) children and adults, respectively. The proportion of children and adults who were seropositive were similar when stratified by sex (Fig. 2F). A similar age was observed between seronegative and seropositive children as well as between seronegative and seropositive adults (Fig 2G).

When examining the relationship between symptomatic infection and antibody response, a higher proportion of seronegative adults were asymptomatic compared to seropositive adults (4/10, 40% vs. 2/32, 6.3%; p=0.02) (Fig. 2H). Symptomatic adults on average had three times more antibodies than asymptomatic adults (median 227.5 IU/mL, IQR 133.7-521.6 vs. median 75.3 IU/mL, IQR 36.9-113.6) and higher viral load (not statistically significant) than asymptomatic adults, although the number of adults who were asymptomatic and seropositive was small (Fig. 2I-J). In contrast, a higher proportion of seropositive children were asymptomatic compared to seronegative children (although not statistically significant) (Fig. 2H), and similar levels of antibodies and viral load were observed in children regardless of whether they had any symptoms (Fig 2I-J). Notably, viral load correlated with antibody levels (Fig. 2K) but not age (Fig. 2L) in both children and adults.

### Cellular immune response in children and adults following SARS-CoV-2 infection

At the convalescent timepoint, seropositive adults had a significantly lower frequency of IgG+ memory B cells, with a corresponding increase in transitional B cells, CD4+ and CD8+ T effector memory (T_EM_) cells compared with uninfected adults. These differences were also observed between seropositive and seronegative adults but were not statistically significant (Fig. 3). There were no differences in IgG+ memory B cells, CD4+ T_EM_ or CD8+ T_EM_ cells in children, however seropositive and seronegative children showed higher levels of transitional B cells compared with uninfected children (Fig. 3). No other differences were observed for any of the other cell populations examined in children or adults (Supplementary Fig. 6). We also compared innate responses during the acute phase in children and adults. We found no differences in innate immune responses for both children and adults based on serostatus, although the number of samples available for this analysis was small (Supplementary Fig. 7).

**Fig. 3:**
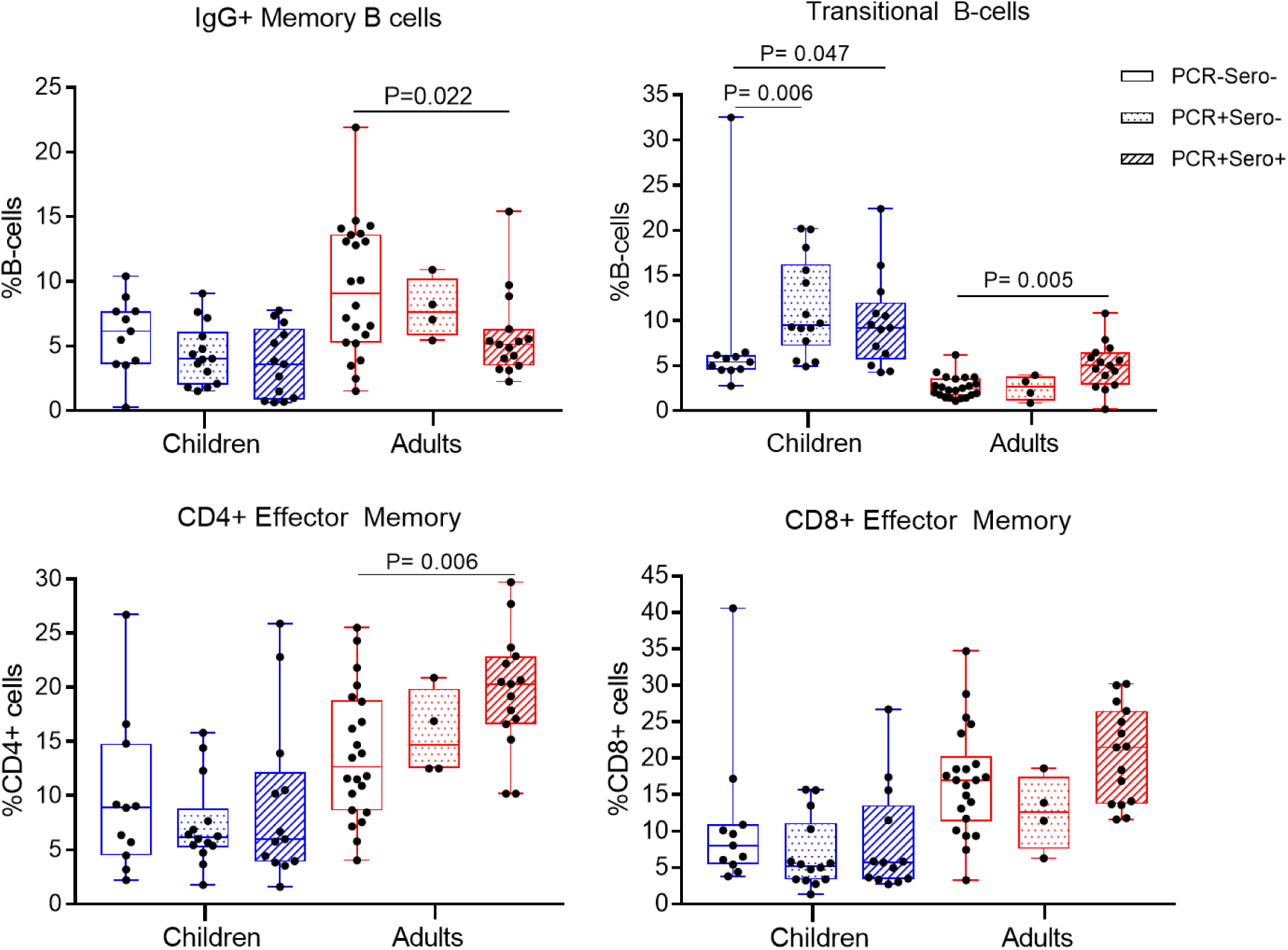
*Ex vivo* cellular immune profile during convalescence period (median day 41) in children (PCR+sero-, N=14; PCR+sero+, N=13) and adults (PCR+sero-, N=4, PCR+sero+, N=15) following SARS-CoV-2 infection; PCR+sero-: dotted box, PCR+sero+: diagonal shaded box. An uninfected control group was included for comparison (PCR-sero-: children, N=11; adults, N=22); clear box. Bars represent median and range.

## Discussion

Our study found that a lower proportion of children with confirmed SARS-CoV-2 infection seroconverted compared with adults despite no difference in viral load. However, SARS-CoV-2 infection in adults generated changes in cellular immune profiles that were most evident in seropositive adults, while this was not observed in children except for transitional B cells. Taken together, our findings provide insights into how children and adults respond differently to the virus. Reduced likelihood of seroconversion may mean that children are less protected against SARS-CoV-2 infections in the long term compared to adults.

Several factors such as age, viral load, sex, comorbidities (including diabetes, cancer and immunosuppression) and disease severity have been found to influence SARS-CoV-2 antibody responses^27-35^. To the best of our knowledge, no data on the proportion of SARS-CoV-2 infected children who seroconvert and the factors impacting this have been reported. A recent study found that 36% of adults with mild COVID-19 did not seroconvert^36^. In comparison to adults who seroconverted, seronegative adults had lower viral load in their respiratory tract and were younger (50 vs 40 years)^36^. The proportion of adults who did not seroconvert were similar to that observed in our study. Other studies in adults have reported variable seroconversion rates between 5 and 25%^30,37-39^. Viral load was similar between children and adults in our cohort, which does not explain why fewer children seroconverted compared with adults. However, our data suggests that Ct value of less than 26 is associated with seroconversion in both children and adults. A similar Ct value threshold of 25 was found to be associated with seroconversion in a previous study^40^. Interestingly, asymptomatic infection was associated with lower seropositivity and antibody levels in adults but not in children, consistent with previous studies in adults^35,41^ and children^8^. This suggests that the host humoral response to SARS-CoV-2 infection in children is different to adults despite similar viral loads and circulating virus variant exposure.

There are several immunological hypotheses as to why children might be less likely to seroconvert. First, antibody profiles (antibody isotypes and subclasses)^12,14,42-44^ and memory B cell populations have been reported to be different between children and adults, but this has mostly been related to disease severity^45,46^. We^25^ and others^12,20,42,43,47^ have previously reported similar SARS-CoV-2 specific IgG antibody levels between children and adults. We however did not measure IgG subclasses (i.e. IgG1 and IgG3) which have also been associated with COVID-19 severity as well as age effects^14,44^. In our study, we observed a decrease in IgG memory B cells among seropositive adults that corresponded with an increase in transitional B cells in SARS-CoV-2-infected children and adults. One explanation for this is that activation of pre-existing memory B cells during SARS-CoV-2 infection leads to increased transitional B cells to compensate for the loss in the B cell compartment^48,49^. Whether transitional B cells play a role in seroconversion remains to be determined.

Second, T cell responses differ between SARS-CoV-2 infected children and adults. A recent study of SARS-CoV-2 T cell responses in children and adults with mild COVID-19 found that infected children had reduced CD4+ T cell effector memory to SARS-CoV-2 proteins compared to infected adults^50^, consistent with our findings although we did not undertake *ex vivo* stimulation experiments. Our data supports the concept that infection may not induce the robust cellular immune responses in children that are necessary for seroconversion as seen in adults^51,52^.

Children are thought to have a more robust innate and/or mucosal immune response to SARS-CoV-2 than adults ^13,15,53-55^. This could explain why children in our study did not appear to trigger the adaptive immune system as well as adults. However, our analysis of innate immune responses in children by serostatus did not reveal any differences, likely due to the small sample size. More efficient innate immunity may also suggest a shorter viral clearance time in seronegative children, but this was not observed in our study. We previously showed that the appearance of mucosal SARS-CoV-2 antibody levels in children was associated with symptom resolution and lack of seroconversion in a family case study^53^. Further analysis of mucosal responses in children are ongoing. Clearly, several factors are likely to contribute to the lack of seroconversion, and more studies are needed to improve our understanding of this response.

Our findings have important implications for protection against SARS-CoV-2 in children. Numerous studies have highlighted the importance of antibodies for protection against SARS-CoV-2. A US study of SARS-CoV-2-infected young adults (18-20 years) reported that SARS-CoV-2-infected seronegative individuals were 80% more likely to be reinfected compared to seropositive individuals. The study also found that low IgG antibody levels in seropositive individuals were associated with reinfection, although seropositive adults had 10-times lower viral load than reinfected seronegative individuals^56^. Therefore, a lack of seroconversion may result in a higher susceptibility to reinfection. This may have important implications on the transmission of SARS-CoV-2 in the community and the public health response.

It is important to note that our findings are based on the ancestral ‘Wuhan’ SAR-CoV-2 virus that was circulating in 2020. The relevance of our findings to current epidemiology where COVID-19 cases in children have been rising due to the SARS-CoV-2 Delta variant^57^ is unclear. Whether lower seroconversion rates in children are also observed following infection with Delta variant is unknown and warrants further research. This variant has been associated with 1000x higher viral load compared with the Wuhan strain so a higher seroconversion rate in children might be expected^58^.

The strengths of our study includes the use of three independent serological assays to examine antibody response against SARS-CoV-2, including a subset of samples that correlated positively with neutralizing antibody assay; these assays have also previously been shown to correlate well with neutralizing antibody assays^25,59,60^. Limitations of this study include the small sample size particularly for the cellular analyses. In addition, our study cohort of mild COVID-19 among children and adults may not be generalizable to other study populations such as older adults or individuals with underlying medical conditions.

This is the first study that documents a higher proportion of children who do not seroconvert compared to adults, despite a similar clinical and virological profile. Seronegative children are at a greater potential risk of reinfection. Our findings have important implications for public health responses in controlling SARS-CoV-2 infection among children and supports COVID-19 vaccination strategies once priority groups have been vaccinated.

## Data Availability

All data produced in the present work are contained in the manuscript

## Acknowledgments

We thank the study participants and families for their involvement in this study. We also acknowledge the MCRI Biobanking service for their help in processing the samples. Funding for the recruitment of participants was provided by the Royal Children’s Hospital Foundation and the Infection and Immunity Theme, MCRI. PVL is supported by NHMRC Career Development Fellowship. D.G.P is supported by a CSL centenary fellowship. K.S. is supported by a National Health and Medical Research Council Investigator grant; her research is supported by funding from the Jack Ma Foundation and the A2 Milk Company. This work is supported by Victorian Government’s Medical Research Operational Infrastructure Support Program. NWC and KS received funding from the National Institute of Health for influenza and COVID-19 research. All other authors reported no conflicts of interests.

