## Supplementary figures for "Reduced seroconversion in children compared to adults with mild COVID-19"

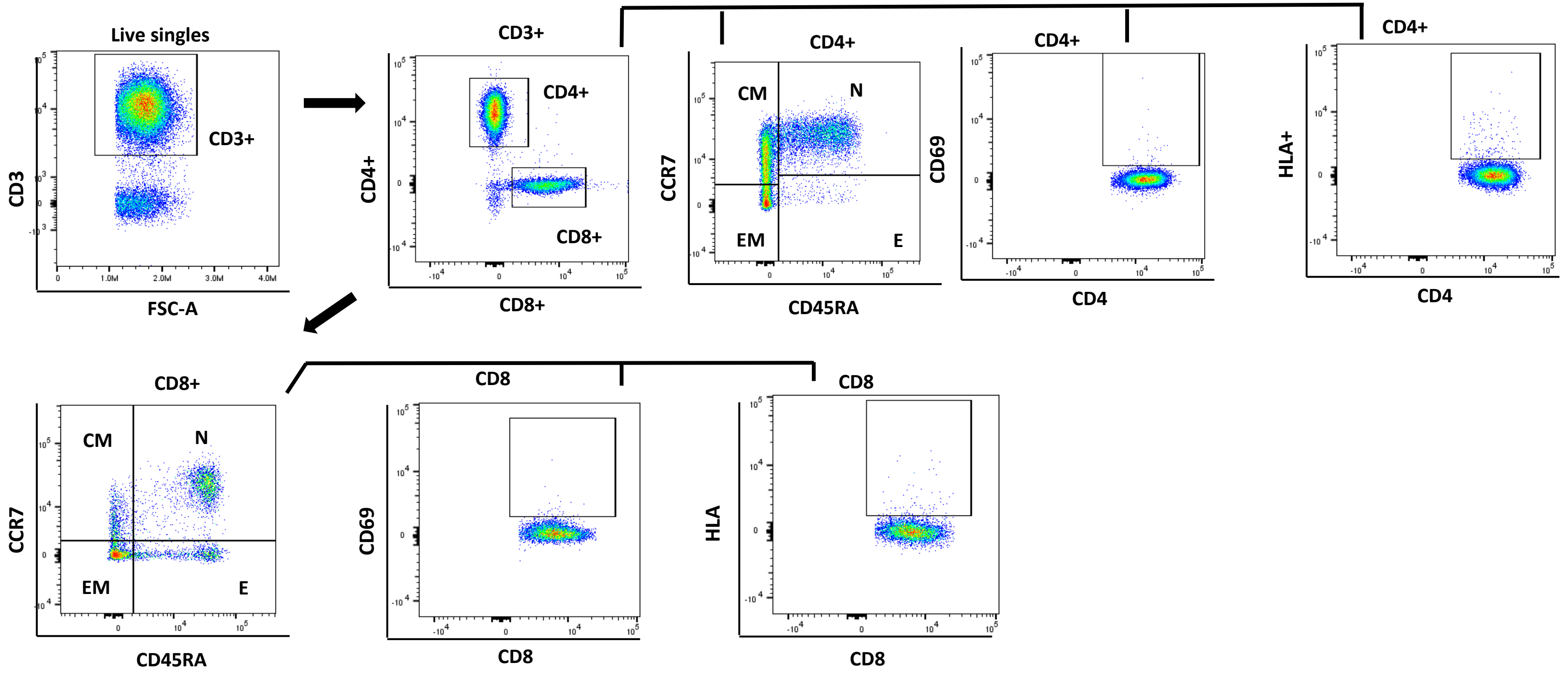


**Supplementary Figure 1: Gating strategy to identify T-cell subsets.** From live single cells, T-cells were identified by positive CD3 expression. CD4+ and CD8+ T-cells were identified from the CD3+ population. CD4+ T-cells and CD8+ T-cells were characterised into naïve (N), effector (E), central memory (CM) and effector memory (EM) based off CCR7 and CD45RA expression. These were CCR7+CD45RA+, CCR7-CD45RA+, CCR7+CD45RA- and CCR7-CD45RA- respectively. CD69+ and HLA+ expression was also characterised on CD4+ and CD8+ T-cells.

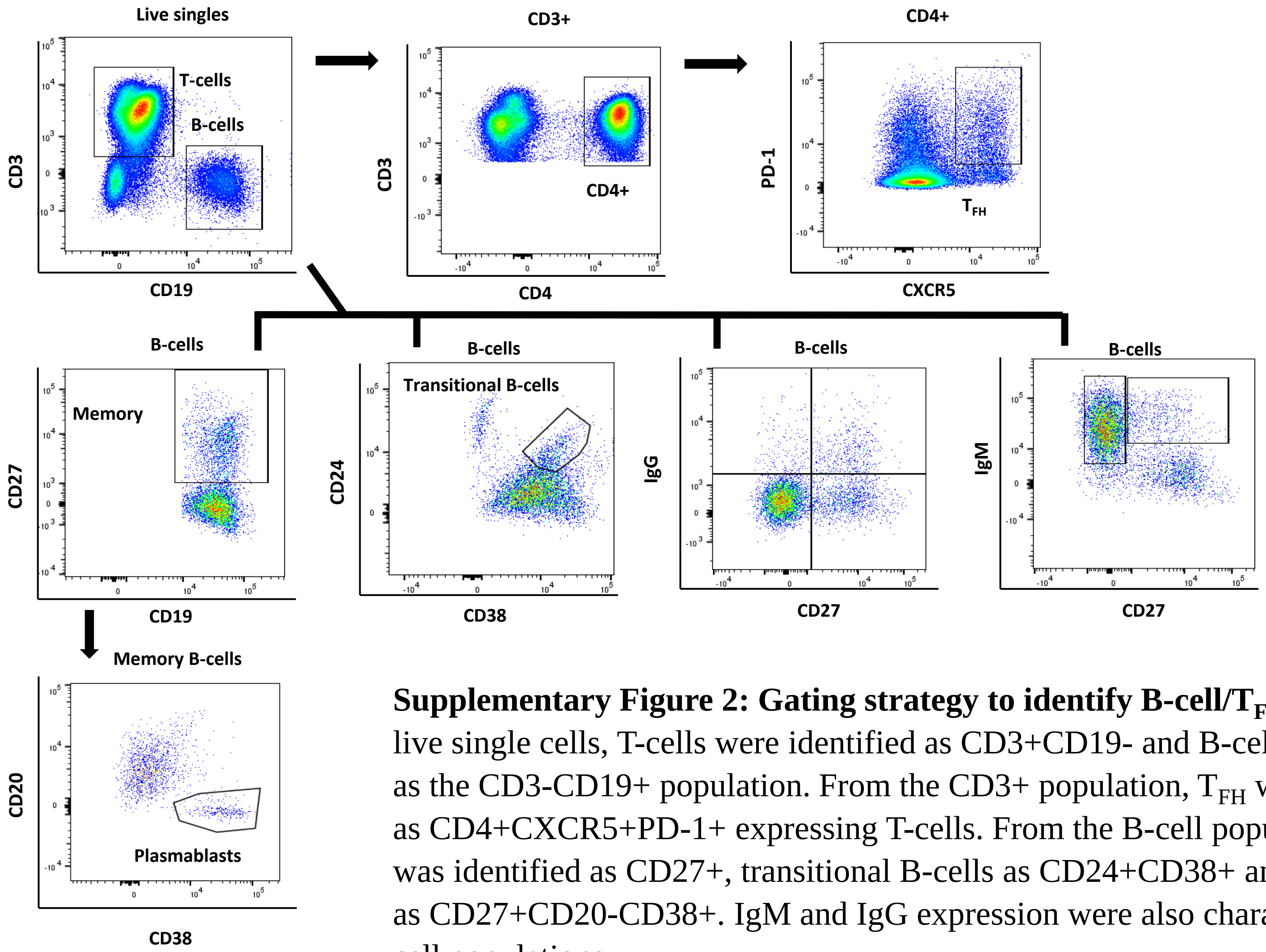


**Supplementary Figure 2: Gating strategy to identify B-cell/T$_{FH}$ subsets.** From live single cells, T-cells were identified as CD3+CD19- and B-cells were identified as the CD3-CD19+ population. From the CD3+ population, T$_{FH}$ was characterised as CD4+CXCR5+PD-1+ expressing T-cells. From the B-cell population, memory was identified as CD27+, transitional B-cells as CD24+CD38+ and plasmablasts as CD27+CD20-CD38+. IgM and IgG expression were also characterised on B-cell populations.

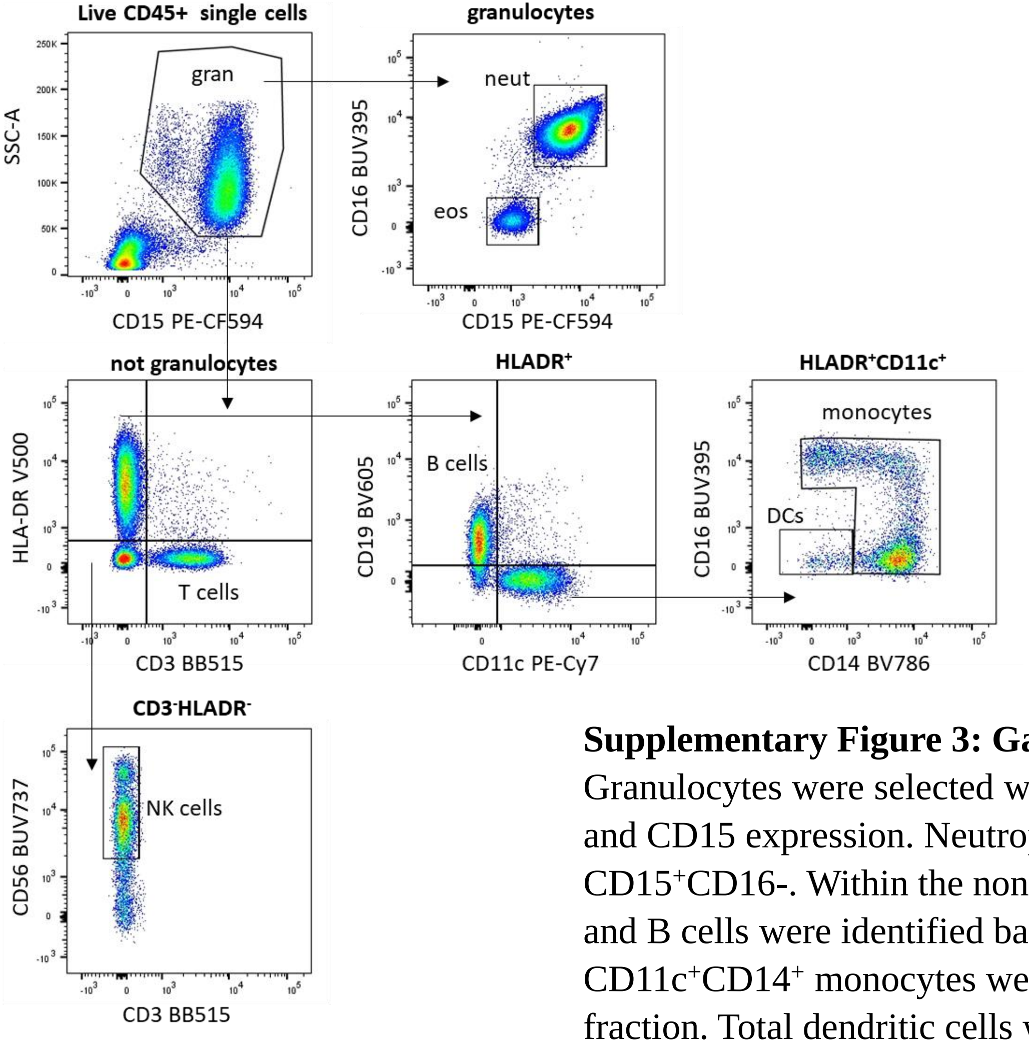


**Supplementary Figure 3: Gating strategy for innate cell populations.** Granulocytes were selected within CD45$^{+}$ leukocytes based on their SSC profile and CD15 expression. Neutrophils were CD15$^{+}$CD16$^{+}$ and eosinophils were CD15$^{+}$CD16-. Within the non-granulocyte fraction, CD3 T cells were identified and B cells were identified based on CD19 and HLA-DR expression. CD11c$^{+}$CD14$^{+}$ monocytes were gated within the non-T cell and non-B cell fraction. Total dendritic cells were HLADR$^{+}$CD11c$^{+}$CD14$^{-}$ and NK cells were HLADR$^{-}$CD3$^{-}$CD56$^{+}$ cells.

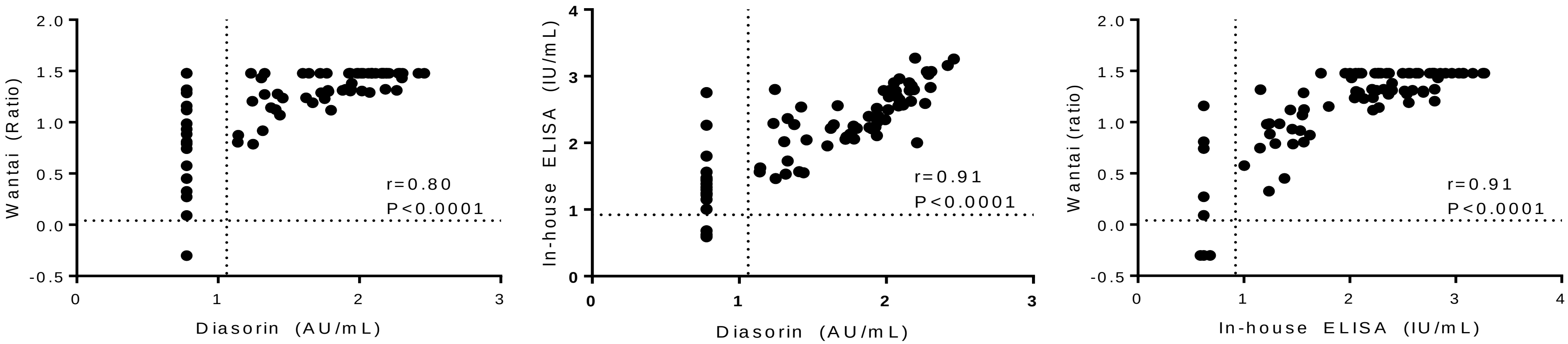


**Supplementary Figure 4:** Correlation analysis of three serological assays using acute and convalescence samples of both children and adults (N=138-145). Antibody levels were log-transformed and analysed using Pearson correlation analyses.

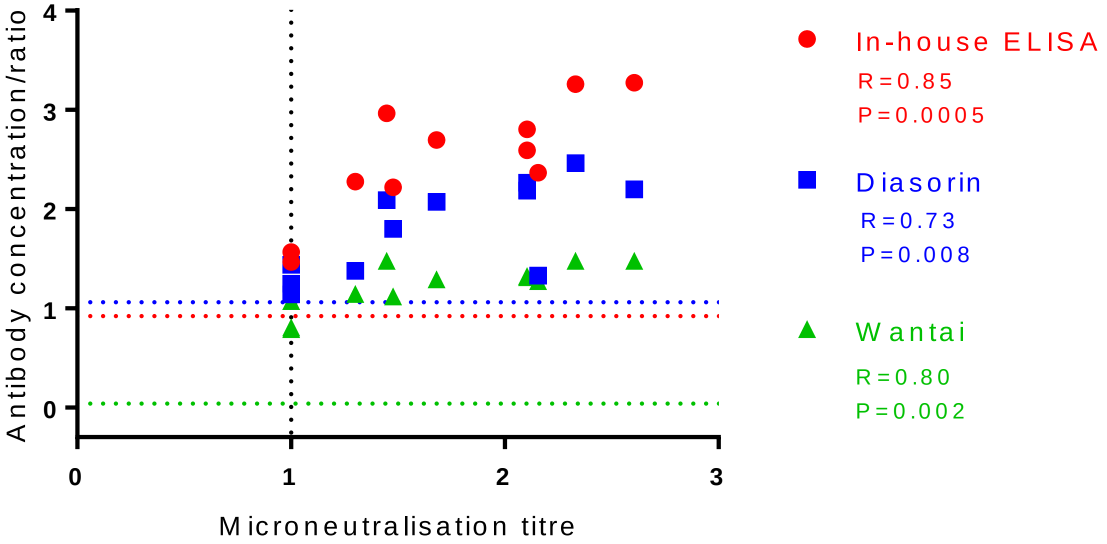


**Supplementary Figure 5:** Correlation analysis of three serological assays against SARS-CoV-2 microneutralisation assay (N=12). Antibody levels were log-transformed and analysed using Pearson correlation analyses. Coloured dotted line represents assay specific cut-off for seropositivity.

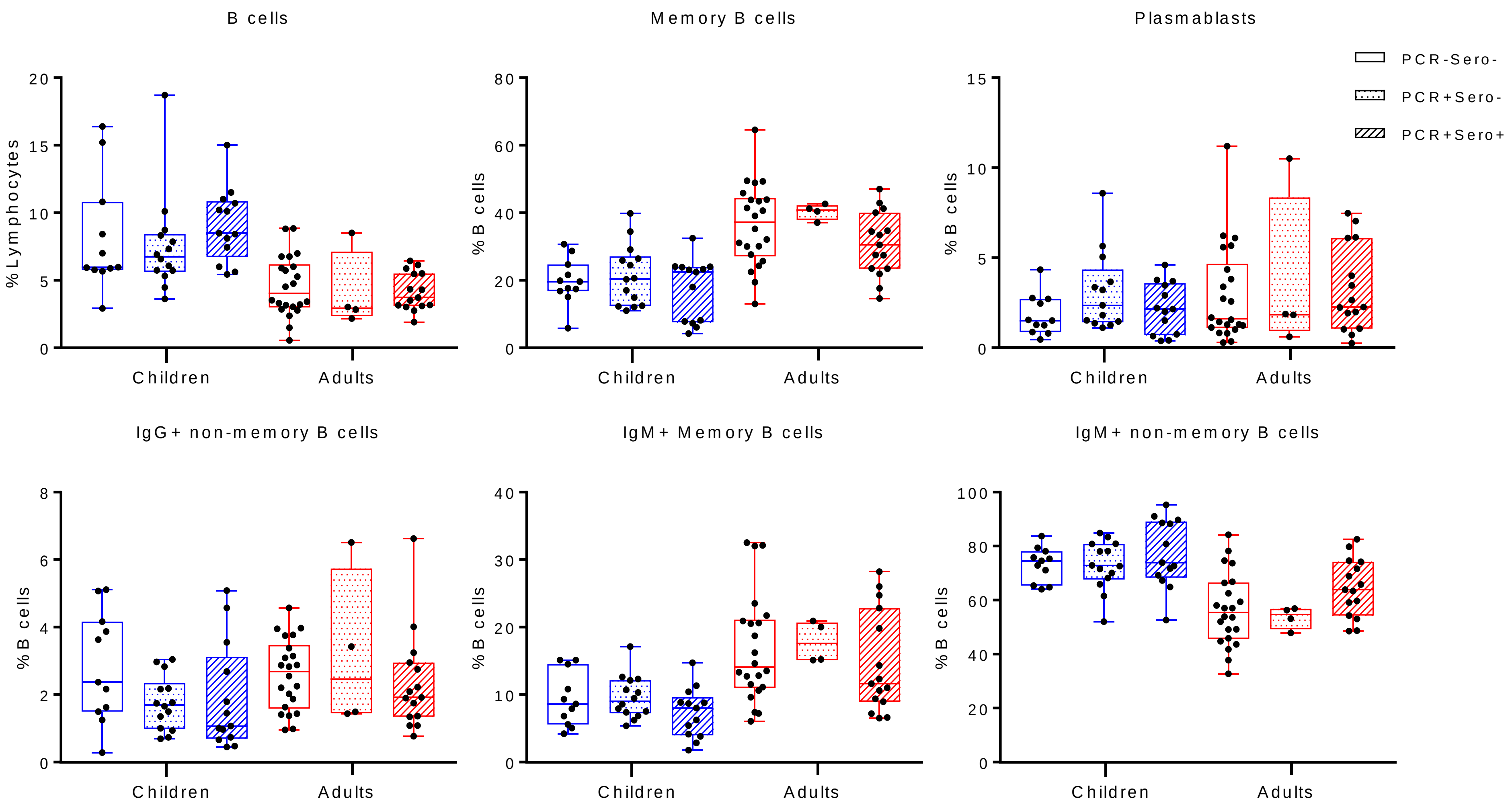


**Supplementary Figure 6a:** Humoral immune cells profile during convalescence period (median day 41) in children (PCR+sero-, N=14; PCR+sero+, N=13) and adults (PCR+sero-, N=4, PCR+sero+, N=15) following SARS-CoV-2 infection. An uninfected control group was included for comparison (PCR-sero-: children,  N=11; adults, N=22). Bars represent median and range.

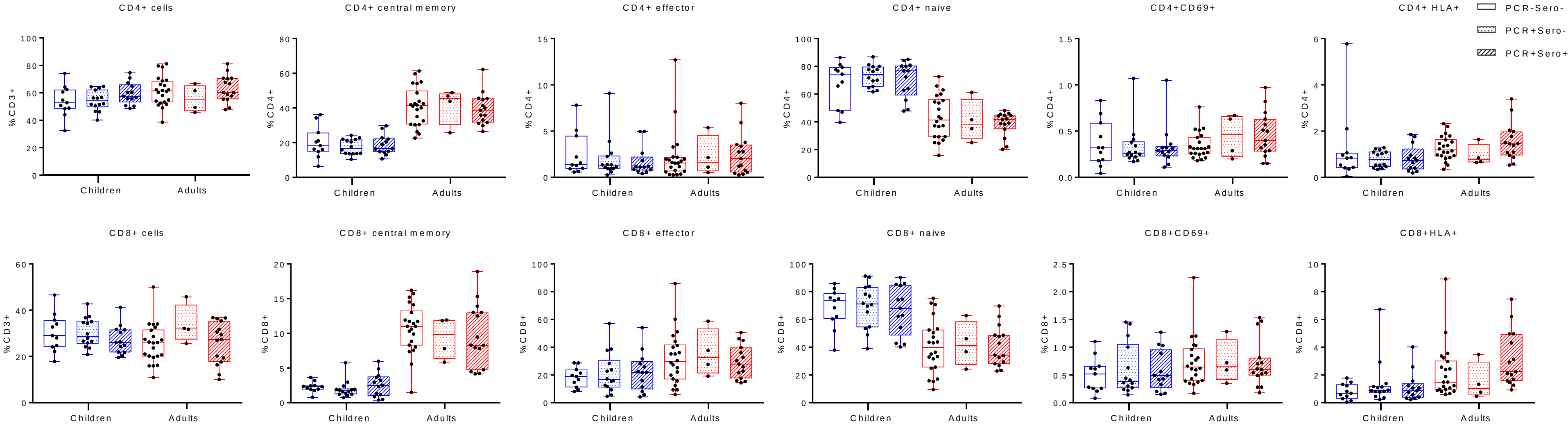


**Supplementary Figure 6b:** Cellular immune profile (T cells) during convalescence period (median day 41) in children (PCR+sero-, N=14; PCR+sero+, N=13) and adults (PCR+sero-, N=4, PCR+sero+, N=15) following SARS-CoV-2 infection. An uninfected control group was included for comparison (PCR-sero-: children,  N=11; adults, N=22). Bars represent median and range.

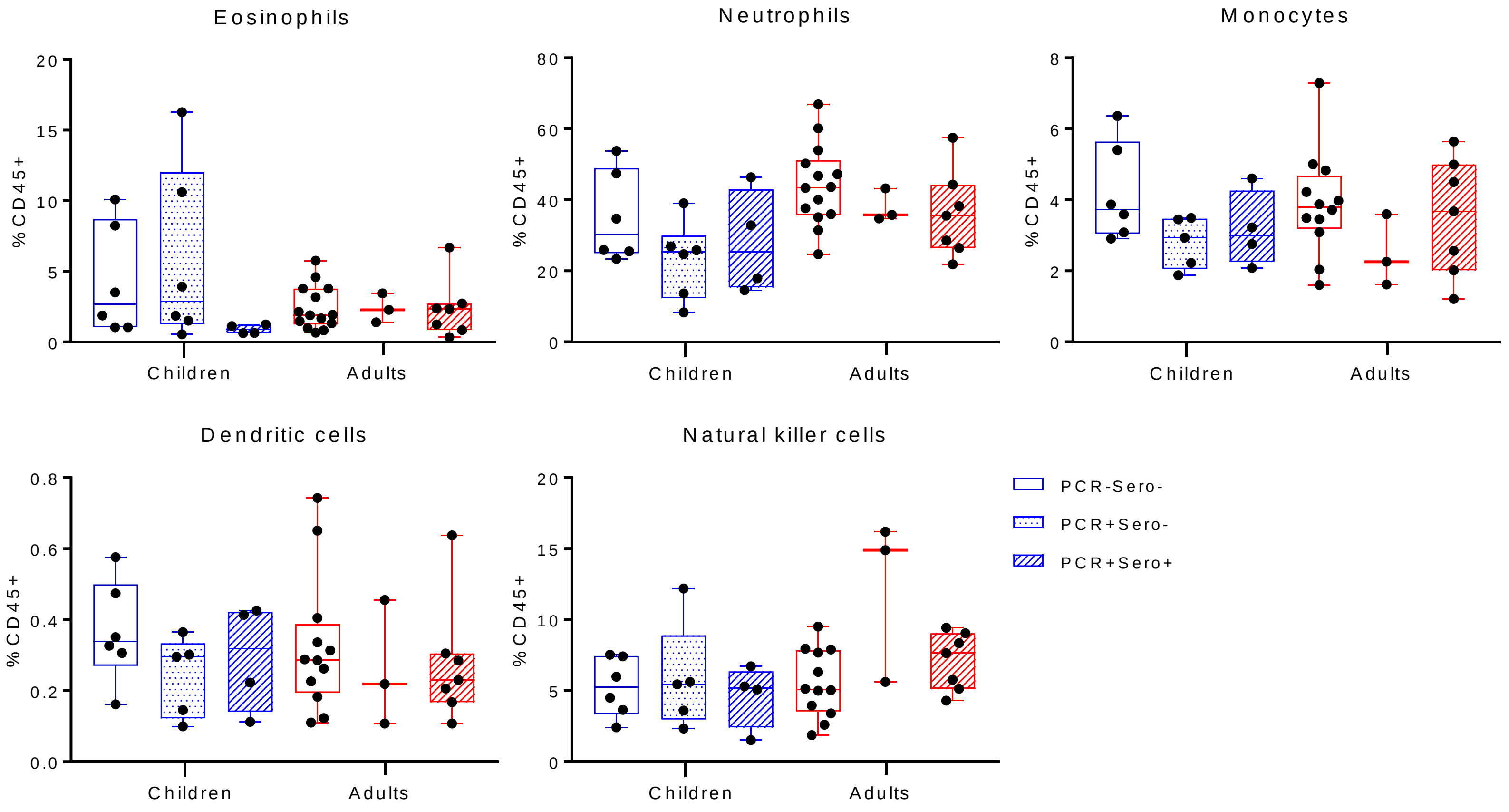


**Supplementary Figure 7:** Innate cell profiles during acute phase (day 7-12) for children (PCR+sero-, N=7; PCR+sero+, N=4) and adults (PCR+sero-, N=3, PCR+sero+, N=8) following SARS-CoV-2 infection. An uninfected control group was included for comparison (PCR-sero-: children, N=6; adults, N=16). Bars represent median and range.
