## Supplementary Tables for "Reduced seroconversion in children compared to adults with mild COVID-19"

**Supplementary Table 1: Antibody cocktail to identify adaptive immune cell populations.**

| Antibody Cocktail 1 | Supplier | Antibody Cocktail 2 | Supplier |
|---|---|---|---|
| CD3-BUV395 | BD Bioscience, San Diego, CA, USA | CD3-Percp/Cy5.5 | BD Bioscience, San Diego, CA, USA |
| CD4-BV421 | BD Bioscience, San Diego, CA, USA | CD4-BV510 | BioLegend, San Diego, USA |
| CD8-BUV805 | BD Bioscience, San Diego, CA, USA | CXCR5-APCR700 | BD Bioscience, San Diego, CA, USA |
| CD45RA-Percp/Cy5.5 | BD Bioscience, San Diego, CA, USA | PD-1-PEcy7 | BD Bioscience, San Diego, CA, USA |
| CCR7-BV785 | BioLegend, San Diego, USA | CD19-BV785 | BioLegend, San Diego, USA |
| CD69-BV650 | BD Bioscience, San Diego, CA, USA | CD20-BV421 | BioLegend, San Diego, USA |
| HLA-APC-H7 | BD Bioscience, San Diego, CA, USA | CD27-BUV737 | BD Bioscience, San Diego, CA, USA |
| Zombie NIR | BioLegend, San Diego, USA | CD19-BV785 | BioLegend, San Diego, USA |
| | | CD38-BUV496 | BD Bioscience, San Diego, CA, USA |
| | | CD24-BV711 | BioLegend, San Diego, USA |
| | | IgG-BV605 | BD Bioscience, San Diego, CA, USA |
| | | IgM-FITC | BioLegend, San Diego, USA |
| | | Zombie NIR | BioLegend, San Diego, USA |

**Supplementary Table 2. Antibody cocktail to identify innate immune cell populations.**

| **Surface Marker** | **Fluorophore** | **Clone** | **Final Dilution** |
|---|---|---|---|
| CD14 | BV786 | M5E2 | 1:50 |
| CD45 | BV711 | HI30 | 1:100 |
| CD56 | BUV737 | NCAM16.2 | 1:100 |
| CD11c | PE-Cy7 | B-ly6 | 1:100 |
| CD3 | BB515 | UCHTI | 1:100 |
| CD15 | PE-CF594 | W6D3 | 1:200 |
| HLA-DR | V500 | G46-6 | 1:200 |
| CD19 | BV605 | SJ25C1 | 1:200 |
| CD16 | BUV395 | 3G8 | 1:400 |
| Live/dead | N-IR | | |

**Supplementary Table 3: Concordance of three serological assays for all samples at convalescent period (Day 41) from the whole household cohort study (95 families)**

| | | **Diasorin** | |
|---|---|---|---|
| | | Positive | Negative |
| **In-house ELISA** | Positive | 56 | 10 |
| | Negative | 3 | 164 |
| | Total | | 233 |
| Agreement | | 0.94 | |

| | | **Diasorin** | |
|---|---|---|---|
| | | Positive | Negative |
| **Wantai** | Positive | 54 | 10 |
| | Negative | 3 | 162 |
| | Total | | 229 |
| Agreement | | 0.94 | |

| | | **Wantai** | |
|---|---|---|---|
| | | Positive | Negative |
| **In-house ELISA** | Positive | 61 | 3 |
| | Negative | 3 | 162 |
| | Total | | 229 |
| Agreement | | 0.97 | |
